# Hypertension perspectives and health behaviors of African youth: The effect of health education and employment

**DOI:** 10.1101/2022.03.23.22272796

**Authors:** M Mhlaba, F Mpondo, LJ Ware

## Abstract

In South Africa between 1998 and 2016, hypertension rates in young adults (15-34 years) more than doubled calling for preventive interventions. However, with many youth unemployed, young adults struggle to prioritize health or implement healthy behaviors. We conducted six focus group discussions comparing hypertension-related beliefs and behaviors between NEET youth (n=20; not in employment, education or training) and previously NEET youth on a health-focused learnership (n=20). While all youth viewed hypertension as life threatening leading to cardiovascular disease or death, especially if left untreated, only youth undertaking health education felt empowered to implement healthy behaviors for disease prevention. In contrast, NEET youth felt hypertension was inevitable and described negative experiences at clinics and fear of lifelong medication use if diagnosed as reasons not to be screened. Our results suggest that engaging NEET youth in culturally appropriate health education programs can motivate preventive health behavior for chronic diseases such as hypertension.

---

Hypertension (HTN) is a leading cause of cardiovascular disease (CVD) mortality; an estimated 17.9 million people died from CVD in 2019 accounting for a third of all global deaths.^1–3^ Over three quarters of these deaths take place in low-to-middle income countries,^4–6^ strongly suggesting a need for urgent health promotion and prevention programs. Sub-Saharan Africa (SSA) has some of the highest rates of HTN globally,^6–10^ particularly among individuals living in urban areas.^7–10^ A recent study including West, East, and Southern African countries reported that adults in Soweto, South Africa were among those who had the highest prevalence of HTN (54.1%).^11^

Additionally, the prevalence of HTN among younger adults is increasing.^1,12,13^ In 2016, the national South African Demographic and Health Study showed that 24% of young adults (aged 15-34years) were diagnosed with HTN compared with only 9% in 1998.^12^ Despite the prevalence doubling in young adults, there remains a general lack of awareness.^14–17^ Due to the fact that HTN often remains asymptomatic,^1,18^ frequently young adults only get diagnosed when being examined for unrelated conditions.^1^ Studies also show that young adults perceive themselves as too “young” to have HTN and therefore generally do not readily go for HTN screening.^18^

Globally young adults have been reported as less integrated into healthcare, less likely to be insured and receive information about lifestyle changes to address HTN.^15,19–21^ In LMICs primary healthcare services often prioritize pediatric care as well as older populations who are more in contact with healthcare facilities.^15,21,22^ Thus, young adults in HIC and LMIC infrequently get screened for HTN compared to other age groups.^20,21^ Additionally, the realities of under-resourced health facilities in Sub-Saharan Africa further exacerbate this situation, widening the service gap for young adults and leading to a lack of engagement in preventive HTN health behavior in this age group.^19,20,23^

Understanding young adults’ perceptions of risk factors and of their own perceived susceptibility is important to design appropriate health promotion and prevention tools, and to know which attitudes and behaviors to target.^6,15^ While there is little qualitative research assessing risk perceptions for HTN among youth globally. Research in South Africa shows that young adults have poor risk perceptions regarding the likelihood and consequences of getting NCDs as well as the usefulness of NCD preventive measures.^24,25^ Identifying social and structural factors that influence health behaviors of young adults is also crucial for designing effective interventions to prevent or manage HTN. For example, one South African study suggests that lower socioeconomic status negatively affects HTN rates in young adults.^26^ Young adults who are not in employment, education or training (NEET) are the most vulnerable in this regard and are most likely to engage in unhealthy behaviors such as substance abuse, poor diet and sedentary lifestyles,^27,28^ as well as have worse mental and physical health.^27^

South Africa currently has one of the highest rates of NEET youth globally,^29^ with a range of youth training initiatives supported by government.^29^ Health education particularly has shown some success in improving health behaviors and modifying CVD risk factors in other regions.^30–33^ Therefore, our aim was to compare HTN related beliefs and health behaviors between NEET youth and previously NEET youth undertaking a health-education training initiative (HETI).

## Methods

### Study setting and participant

The study was conducted from August to October 2021 at a youth development center in Soweto, Johannesburg. Soweto is a densely populated, historically disadvantaged, urban township where the population is predominantly black with a small percentage classifying themselves as middle-class and the majority having a low socioeconomic status.^34^ More than half of the youth in Soweto are unemployed with low education levels and have access to few opportunities with limited work experience or skills training. ^35,36^ The current COVID-19 pandemic has worsened the socio-economic situation for many households.^35^ Additionally, the area houses many taverns and informal convenience stores that sell alcohol, single stick cigarettes and energy dense food at low prices, promoting unhealthy lifestyles and encouraging pro-HTN risk behaviors, ^37,38^ This is challenging particularly in a community with a high concentration of NCDs (i.e., diabetes cases and mental health issues) that greatly impact the quality of life.^39^

Purposeful criterion sampling was used to select NEET youth (NEET group) and previously NEET youth enrolled in a 12-month health-education training initiative (HETI group). All participants were youth (aged 18-34 years) registered at the youth development center for employment and training opportunities. Recruitment took place through a communication platform (WhatsApp) at the center with those who showed interest to participate invited to attend the center to obtain informed consent.

### Data collection

Focus-group discussions (FGD) took place in a private room with six to eight participants in each and each FGD lasted just over an hour. Strict adherence to all COVID-19 protocols was maintained throughout the discussions. Discussions were conducted using a semi-structured topic guide (Table 1) designed around the constructs of the Health Belief Model (HBM; perceived susceptibility, perceived severity, perceived benefits, perceived barriers, self-efficacy, cues to action and individual health behavior).^40^ Self-efficacy was further added as a construct as it has been added to the HBM on many occasions since the late 1970’s,^40^ and was included in the model for purposes of this study. The HBM is commonly applied to health education and promotion studies,^25^ including studies conducted in similar urban African contexts,^24,25^ such as one study evaluating age-group differences in NCD risk perceptions.^24^ The FGD facilitators included a post-graduate level researcher and an observer who were both multilingual. All FGDs were audio recorded and conducted in English, with participants encouraged to use local languages to express themselves more freely if needed. Audio recordings were transcribed and translated into English, and then crosschecked by a third researcher who was fluent in the languages used.

**Table 1.** Focus Group Discussion Topic Guide and related Health Belief Model (HBM) Constructs.

| Question | HBM Construct |
| --- | --- |
| 1. What do you know about high BP/HTN? |  |
| 2. What are your personal views/ feelings about high BP/HTN? |  |
| 3. How severe do you think HTN can get? Probe: What do you think are the consequences to a person's health? | Perceived severity |
| 4. Who in your opinion do you think is most likely to get high BP? | Perceived susceptibility |
| 5. If you were told that you are very likely going to develop HTN, what would your response be? Probe: Do you think you are at risk? Probe: what do people who are at risk look like or how do they behave? (e.g., things that they do in their everyday life) | Perceived susceptibility and severity |
| 6. How important do you think it is to know if you're at risk of getting HTN or to monitor your BP? | Perceived susceptibility and severity |
| 7. How important do you think it is to go to a health center for the diagnosis and treatment of HTN? Probe: Do you think this is always necessary? | Perceived benefits |
| 8. What do you think are other lifestyle changes that you can make to prevent or manage HTN besides taking medication? Probe: which of these lifestyle changes do you think is most important in preventing or managing HTN? | Perceived benefits |
| 9. How likely are you to agree that these are reasonable lifestyle changes to make? | Perceived benefits |
| 10. How do you think engaging in regular exercise or healthier diet can prevent/ manage HTN? | Perceived benefits |
| 11. [HETI group only]: What is the likelihood that you would have gone for a BP check, if you had not been screened by the [name of HETI program]? Probe: Is this a motivation to continue getting regular check-ups?<br>[NEET group only]: What is the likelihood that you would go to the clinic or any health facility for a BP check? Probe: Is this a motivation to continue getting regular check-ups? | Cues to action |
| 12. How did you feel before taking your BP measurements? |  |
| 13. How did you feel once you got your BP results? |  |
| 14. What changes would you like to make/ have you made after taking your BP measurements? Probe: What motivated you to make these changes? Probe: How confident are you that you will stick to these changes? | Cues to action and Self-efficacy |
| 15. How do you feel about taking BP measurements again? |  |
| 16. How often would you feel comfortable taking your BP measurements? | Self-efficacy |
| 17. Would you ever consider taking HTN medication if you were diagnosed with HTN? Probe: What would prevent you from taking HTN medication? Probe: What would prevent you from accessing treatment for HTN? | Perceived barriers |
| 18. What would prevent you from engaging with healthy behavior or changing your lifestyle to prevent HTN? | Perceived barriers |

### Ethics

Ethical committee approval for the study was obtained from the University of Witwatersrand Human Research Ethics Committee (Medical) [reference M210549]. All participants invited to take part agreed and all gave informed consent to participate in the study and to have their responses audio recorded.

### Analysis

Transcripts were analyzed using deductive thematic analysis informed by the HBM constructs. In the analyses, Braun and Clarke’s six-phase analytical process was undertaken, involving (1) familiarization with the data; (2) generating initial succinct codes; (3) identifying themes; (4) reviewing themes; (5) defining and naming themes expressed in relation to the data set and research question; and (6) producing the report.^41^ The first author conducted the initial coding process and created a coding frame. The code extraction, coding frame and interpretation of codes and themes were reviewed with the second and third author, with disagreements resolved through discussion. Upon agreement, the first author summarized the findings.

## Results

In the results, we present the perceptions of the young adults under each of the HBM constructs, identifying similarities and differences between the NEET and HETI group for each construct.

### Perceived susceptibility

Within this theme, the two groups differed in their perceptions and had no common ideas on susceptibility emerging. For example, the two groups viewed the presence or absence of physical symptoms of high blood pressure (BP) differently. The HETI group were aware that the absence of symptoms did not imply low susceptibility to HTN. While the NEET group perceived a lack of HTN-related symptoms to mean they were not susceptible and a reason for not engaging in BP screening. The HETI group described how their perception of risk had changed through taking part in the health education and training initiative.

*PT 6 - F; NEET group: I would know if something is wrong with me. So only then I would panic and go (screen for HTN), but for now, I don’t think its important*.

*PT 3 – F; HETI group: … I was twenty-four years old, I was told I could get it (HTN) […]so I knew but I didn’t do anything cause I wasn’t feeling anything. Fast forward to now, I know better*.

The two groups also viewed a HTN diagnosis in relation to age differently. While the HETI group expressed that all age groups are susceptible to HTN, the majority of the NEET group perceived HTN as an older person’s illness, and they viewed a HTN diagnosis as being inevitable at an older age.

*PT 3 –F; HETI group: Anyone is most likely to get it (HTN) if maybe they don’t have like a good lifestyle*.

*PT 6-M; NEET group: I always thought it was for old people this thing (HTN)*

*PT 5-M; NEET group: I feel like (HTN) it’s a norm cause eventually the stress of life as you grow up, we are gonna have it*.

The HETI group were aware of their personal risk for HTN and were motivated to engage in preventive health action. In contrast, the NEET group were also aware of increased risk but did not feel inclined to act on this.

*PT 4 – F; HETI group: …for me I think I am at risk of getting HTN because I am obese obviously; I am not free because now I have to watch my weight and also check on my high blood regularly*.

*PT 7 – M; NEET group: Well to be honest […] I do have some information about it (HTN)…the thing is I’m not concerned about it […] both sides of my family got high blood, my mother and my father*.

### Perceived severity

Participants from both groups recognized that HTN is life threatening and related to other cardiovascular conditions, although the NEET group considered HTN as exposing one to additional health issues such as gout.

*PT 6 – F; NEET group: Yes, it leads to death cause my grandma recently passed on. PT3-F; HETI group: …if it is not treated it can lead to health issues and it is very likely for one to get a stroke*.

*PT 3 – M; NEET group: High blood can also make someone to have diabetes*.

### Perceived Barriers

Both groups identified a lack of support as a major barrier for adopting preventive health behaviors. For the HETI group this was primarily in relation to support from family for eating a healthy diet. For the NEET group, it was support from healthcare professionals, they felt mistreated by clinic staff and saw primary healthcare service provision as inefficient and inadequate and chose not to engage with primary healthcare facilities.

*PT 6 – F; HETI group: … I can’t change my diet alone cause [name] at home is okay, she has her pap*^*1*^ *and I can’t eat pap. It’s not easy to change your diet alone, we need to get support from our families as well*.

*PT 4 – M; NEET group: “You go for an injection; she (the sister) will look and say you are here to waste time. So, it’s so boring for us (young people), whether it’s to go and check HTN or anything else you cannot go because of the treatment that you get there”*.

*PT 7 – M; NEET group: “…maybe it’s your day to take the treatment then you go to clinic and the say […] we don’t have treatment today, come next week”*.

The two groups had different viewpoints regarding BP measurement. Many from the NEET groups had not checked their BP and did not want to be screened or know their HTN status. They described the knowledge of having HTN as a trigger for stress, and that in itself would lead to deteriorated health. In contrast, the HETI group saw checking BP to give only benefits.

*PT 1 - F; NEET group: “… with many diseases, if you don’t know, you continue with your life as if it’s normal. But once you go there to the health center and they say you have this. Now it’s going to be in my mind all over and then I’m gonna be stressing and stressing and I’m gonna end up dying sooner*.*”*

Both groups presented feelings of vulnerability and fear around embarking on lifelong HTN medication should this be necessary, this also demonstrated low levels of self-efficacy for this behavior. Youth feared that medication would expose them to other illnesses or side effects, which would ultimately lead to a deteriorated health condition.

*PT 2 – M; NEET group: “I just personally feel that medication is not healthy, it exposes you to other illnesses”*.

*PT 1-M; HETI group: Just the thought of taking a pill everyday […], it’s just a no no, and the side effects apparently*.

Both groups also viewed taking medication to manage HTN as a chore. Many referred to it as a lifetime obligation, where the idea of taking medication every day, over many years was seen as a burden, especially if starting at a younger age. Young adults were reluctant to take medication as a treatment option, expressing that they would rather engage in alternative measures to reduce BP.

*PT 2 – F; HETI group: “Now…no, I’m still young (to start taking medication). I’d rather stop like alcohol […] before medication. Cause it’s not a six months treatment […] it’s a lifestyle, lifetime thing”*.

*PT 4 – F; NEET group: “… last time at the clinic I saw, they were giving them, […] three or four packets, […] it’s too much, you have to take a lot of pills”*.

### Perceived benefits

Groups had different views regarding the benefit of healthy behavior for HTN prevention. The HETI group perceived screening and knowing their BP status as a benefit that empowered them to prevent disease and prolong their lives.

*PT 1 – F; HETI group: “…it’s important to know your status and go for health checks, also because when you are pre-hypertensive it’s still entirely dependent on you to manage it as opposed to when you are now diagnosed with HTN, […] you are dependent on the medication to manage the HTN for you”*

*PT 5 – F; HETI group: “… I think it’s (BP screening) important because it determines how long you gonna live, because if you don’t take care of yourself obviously, you’re reducing your lifespan*.*”*

However, the NEET group did describe engaging in some preventive health behavior but the benefit of this was perceived as stress management and not necessarily to reduce CVD risk.

*PT 4-M; NEET group: Right now I am someone who gyms […] and then everything including stress finishes because of going to the gym*.

### Self-efficacy

In both groups, young adults’ confidence in their ability to engage in health behaviors to prevent HTN was influenced by their level of motivation. For the HETI groups, self-efficacy was bolstered by regular BP checks through daily access to BP monitors. This provided motivation through feedback. However, the NEET group, without access to such equipment or health checks and no desire to engage in preventive behavior, had lower confidence levels.

*PT4 –M; HETI group: “I think we have a good motivation currently to actually make sure that we at least try to initiate the change in our lifestyles because of the health checks that we’ve been doing; that is a motivation”*.

Both groups had low confidence levels in their ability to manage their diet. NEET group also mentioned using food to manage stress levels, a behavior they felt would be challenging to manage or change.

*PT 1-F; HETI group: “I love my food [*…*] so those kinds of things are still kind of difficult to change, even though I try to you know manage and limit some things”*.

*PT 5-M; NEET group: … food (is difficult to manage) because the minute I start stressing the first thing I think of is the food”*.

### Cues to Action

Participants described how they were exposed to health messages in their environment, which raised awareness about health and, at least for the HETI group, appeared to act as cues to action. For the HETI group, health messages came through their health education. For the NEET group, health messages came from clinic health checks or community-based health campaigns, though neither were focused on HTN.

*PT2 –F; HETI group: “The last time I spoke to (name of health educator) she told me that instead of getting off at my stop at the taxis; I should maybe get off two stops before my stop and walk, so I think I walk more than before now”*.

*PT 3 – F; NEET group: “I do take […] contraceptives, so I have to test (for BP)”*.

*PT 3-M NEET group: “The first time I tested for HIV, it wasn’t at the clinic, it was at those tents that they put like maybe at the mall, for me it’s convenient”*.

However, this health messaging was insufficient in the NEET group to drive action, and many said they would need to experience a symptom of HTN to consider going for BP measurements.

*PT 7-F; NEET group: “If I felt that there are symptoms or […] that I’m not feeling well then I would go (get checked) but if I’m not feeling anything or I feel healthy then like no I wouldn’t*.*”*

### Individual health behaviour

Lifestyle modification was a central feature of discussions around individual’s health behavior. The HETI group attributed their change in behavior to access to HTN health education and frequent BP checks. As a result, they expressed that they were more vulnerable to HTN and mindful of its consequences. They perceived more benefits in healthy living to prevent HTN, felt more confident and motivated to control their BP and responded more positively to health action recommendations. This resulted in more proactive health behaviors such as being more physically active, frequent self-monitoring, and improved health outcomes.

*PT 6 – M; HETI group: “Tracking my BP (referring to changes made). I actually bought the machine at home (BP machine), we have a day where we actually check”*.

*PT3 – F; HETI group: “I also walk more now […]. Even my BP is low now”*.

The NEET group gave no evidence that they had or were inclined to implement any lifestyle change to prevent problems with BP. Those who engaged in health behavior such as physical activity did so for other reasons. This group had a reactive approach to HTN health behavior as something they can only engage with in the event that they experience symptoms or have a HTN diagnosis.

*PT 6 – F; NEET group: “I would honestly ignore you (If you told me I had HTN). I would honestly let it slide. It won’t change my lifestyle completely […] I don’t want to change my lifestyle”*.

## Discussion

The aim of this study conducted among young adults from a historically disadvantaged township in South Africa was to explore perceptions of hypertension risk and preventive health behaviors, using a qualitative study centered on the health belief model (HBM) and comparing NEET youth (not in employment, education or training) and previously NEET youth on a health education training initiative (HETI). Our study found that health education influenced all constructs of the HBM and while perceived barriers remained for the HETI group, they were proactively engaged in health behaviors to prevent hypertension. In contrast, in the NEET youth (group without health education), perceived barriers related to healthcare service provision and low perceived susceptibility and resulted in no engagement with hypertension preventive behavior. This was despite NEET youth being aware of the severity of HTN and holding perceptions that it was an inevitable outcome for them as they aged. A detailed interpretation of these findings, assessing both groups, existing literature as well as implications for hypertension preventive health behavior and health promotion are discussed below.

There is scant data on how youth understand their risk and susceptibility to NCDs, globally. However, two previous South African studies showed how a lack of perceived risk for developing NCD’s was linked to young adults’ underestimation of the consequences of unhealthy behavior.^24,25^The young adults in these studies were also shown to have limited awareness of the benefits associated with health behavior change.^24,25^ Our study demonstrates that health education can increase both awareness and personal relevance for youth, thus promoting the adoption of healthy behaviors. The fact that our study demonstrates that the young adults found personal relevance is critical because studies focused on health behavior change argue that for a recommended health action to have lasting effects, individuals must find personal relevance.^42–45^

Our study showed that NEET young adults were not engaging with current health services. Studies on a local and global scale have identified barriers to health service accessibility for young people as lack of insurance, limited clinic time, financial barriers,^15,46^ and similar to our study perceived negative attitudes of health providers and perceived poor staff competencies.^19,20^ Additionally, research has also demonstrated that primary healthcare (PHCs) facilities in SSA are under-resourced, with limited human and medical resources, hindering youth-friendly healthcare provider policies. ^19,20,23^ This in turn hinders screening, preventive and control measures for chronic illness in this age group globally. Our work suggests that community-based services may be able to bridge the gap between the primary healthcare inadequacies and the youth accessing screening or general care services, especially to support the prevention of NCDs. These services have been useful in supporting hypertension and diabetes awareness and management in other age groups in South Africa.^47^This is especially relevant in low-to-middle income countries like South Africa where PHC is under-resourced and overburdened.^19,20,23^

Our study also shows that NEET youth wanted to prevent mental illness and they used that as a motivator for behavior change; the NEET youth expressed engaging in regular physical activity for stress reduction, however it was not necessarily for blood pressure management. Physical activity could be used in future interventions as a behavior change objective; community-based services could encourage young adults to engage in physical exercise to promote ‘double benefits’ (i.e., short term stress reduction and longer term decreased CVD risk), thereby increasing perceived benefits in this group. Physical activity and/or stress reduction interventions are well known globally to reduce BP and CVD risk.^48–52^ Such interventions may be particularly salient for NEET youth, who have been reported as highly vulnerable to stress as a result of their NEET status.^27,53^

Our results also showed that young adults generally fear embarking on lifelong medication use. This is consistent with other research and suggests that medication adherence in young adults is generally low,^18,54,55^ and medication side-effects are the main barrier to adherence in young adults with HTN.^18^ Additionally, individuals who default on taking antihypertensive medication perceive no immediate negative health consequences,^18^ potentially reinforcing the decision to not take treatment. The unfriendly youth services at PHC facilities as described by NEET youth, have previously been shown to be a barrier to medication adherence for other chronic illnesses among South African youth.^20^ Our findings suggest that health education is insufficient to change medication hesitancies and young adults specifically may be highly treatment resistant in the event of a HTN diagnosis. While this again highlights the need for more preventive action, further research is needed to understand how best to support the needs of young adults that require chronic medication and through which services, especially considering the growing HTN burden at younger ages in low-middle income countries.^56^

Studies using the HBM to evaluate the significance of structured educational programmes on NCD risk preventive behavior reported improvements in participants perceived benefit and attitude towards a healthy lifestyle, ^57,58^ as was the case for the HETI group in our study. While studies reporting the positive effect of health education on health behavior are prevalent in older people.^30–32,57,58^ Other research suggests younger adults on prolonged health education interventions are more prone to adopt healthier lifestyles.^59^ While supportive methods (i.e. phone calls, message reminders and reading materials) are suggested as crucial to reinforce positive hypertension health behavior in short-term educational interventions.^60^ This has been suggested as insufficient for hypertensive young adults who consider such methods as an invasion of their privacy and increasing the likelihood of exposure and stigmatisation from peers.^18^ These young adults also reported discarding educational materials.^18^ Further work is needed to understand optimal HETI duration and necessary components required to promote and maintain medication adherence in young adults. Furthermore, with the right education, awareness and community screening, youth may be motivated to engage in preventive behaviors if they are likely to need medication soon.

Our results suggest that investments into health education learnerships, would result in double duty benefits, addressing low education and employment levels in South Africa’s young adults, where the NEET rate is among the highest globally.^29^ In a context where healthy behaviors among young adults are frequently unobtainable in the absence of secure employment and career prospects.^61^ Moreover, where NEET young adults are more vulnerable to risky health behavior,^27^ and where poor health has been reported to further give rise to a NEET status.^62^ Such interventions would shift young adults focus to their health awareness and behavior. Ensuring they make better health choices in the short term, aiding to reductions in NCD’s in the long term. A report showcasing youth education interventions in four African countries showed that only one out of forty-seven considered health, promoting young people’s access to sexual and reproductive health services.^63^ Youth education learnerships promoting access to broader NCD services are urgently needed, as the number of young adults with hypertension increase globally.^1,12,13^

### Limitations and strengths

Utilising the HBM provided a useful framework for investigation but may have also created some limitations. While the HBM has been used many times to explore perceptions around health in young adults in similar contexts to our own.^24,25^ it is possible that a further construct representing perceived importance may be needed to better understand health behavior outside of a health education context. Our findings suggest that this may be the case, as others have also previously suggested.^45^ A further limitation is that this study only included young adults from one historically disadvantaged location. However, our findings are supported by previous research from across countries showing similar experience among young adults in relation to NCD risk perceptions and health behavior.^15,18,24,25^

One strength of this work is the investigation of how socio-economic status affects hypertension health beliefs and behaviors in youth who are not diagnosed with hypertension. This assessment is critical in a country where two out of three young adults are unemployed.^35^ Additionally, while a number of studies have analyzed risk factors, perceptions and self-care behaviors among young people already diagnosed with HTN,^12,18,54,55,64,65^ this study focused on preventive measures in young adults without a known diagnosis in which prevention efforts may be most beneficial.

## Conclusion

Our study set out to evaluate hypertension perceptions and health behaviors in young adults not in employment, education or training (NEET) in Soweto, South Africa, comparing these with perceptions and behaviors of young adults engaged in a health education and training initiative. Our findings suggest that health education proactively engaged youth in health behaviors to prevent hypertension. In contrast, NEET youth reported no engagement with or intention for hypertension preventive behavior despite the perception that hypertension was severe and an inevitable outcome for them as they aged. Given the governments drive for youth employment and education initiatives, those that focus on health may show multiple benefits for the future of South Africa’s young adults.

## Data Availability

All data produced in the present work are contained in the manuscript

## Footnotes

1 Pap is a traditional South African staple food made from softly grounded maize.

